# The Immune-Buffer COVID-19 Exit Strategy that Protects the Elderly

**DOI:** 10.1101/2020.09.12.20193094

**Authors:** Vered Rom-Kedar, Omer Yaniv, Roy Malka, Ehud Shapiro

## Abstract

September 12, 2020

COVID-19 is a viral respiratory illness, caused by the SARS-CoV-2 virus with frequent symptoms of fever and shortness of breath [1]. COVID-19 has a high mortality rate among elders. The virus has spread world-wide, leading to shut-down of many countries around the globe with the aim of stopping the spread of the disease. To date, there are uncertainties regarding the main factors in the disease spread, so sever social distancing measures and broad testing are required in order to protect the population at risk. With the increasing spread of the virus, there is growing fraction of the general population that may be immune to COVID-19, following infection. This immunised cohort can be uncovered via large-scale screening for the SARS-CoV-2 (Corona) virus and/or its antibodies. We propose that this immune cohort be deployed as a buffer between the general population and the population most at risk from the disease. Here we show that under a broad range of realistic scenarios deploying such an immunized buffer between the general population and the population at risk may lead to a dramatic reduction in the number of deaths from the disease. This provides an impetus for: screening for the SARS-CoV-2 virus and/or its antibodies on the largest scale possible, and organizing at the family, community, national and international levels to protect vulnerable populations by deploying immunized buffers between them and the general population wherever possible.

**Declarations of interest:** none

## 1 Introduction

An urgent need of humanity in the face of the COVID-19 pandemic is to buy time, until treatments and vaccines to the disease become broadly available. The question arises how to best organize the conduct of families, communities and nations in order to minimize morbidity and mortality until treatments and vaccines are available.

A salient characteristic of COVID-19 is that specific sub-populations suffer much higher rates of severe illness requiring hospitalization and intensive care, as well mortality, relative to others. These include especially older age-groups[2, 3], as well as individuals suffering from background illness: hypertension, obesity, diabetes and coronary heart disease [4, 5]. Data from China shows that 81% of fatalities are in the 60+ age group, and in Italy 96.5% of fatalities are in the 60+ age group[2]. Protecting these groups will thus be a central goal of any strategy for alleviating the burden of the current pandemic.

Despite efforts to suppress transmission, it is likely that a significant fraction of the population will eventually be infected, and a minute fraction of these may suffer from re-infection (as of August 2020, out of above 17 million recovered patients there are only 3 verified re-infection cases). While a majority of young and healthy individuals will recover following mild illness, or even undergo asymptomatic infection, the high likelihood of widespread transmission underscores the need to protect the vulnerable from infection. At present this can be achieved only by isolating vulnerable individuals from potential sources of transmission. However, it is precisely these vulnerable individuals who are often dependent on continual care and support from family members, medical staff, nursing home staff, and others. Nursing homes for the elderly have tragically become hot-spots of infection [6, 7]. By their very nature2018;caretakers’, on which their well-being depends.

Here we propose an approach, termed Immune Buffer Strategy (IBS), aimed at protecting vulnerable populations while easing restrictions on the general population and on the interactions of the vulnerable individuals with some of their care takers. The key idea is to make use of able people who have been infected and recovered, as an active buffer between the population at risk and the population at large.

Depending on the governance mechanism in the country, or the social norms in the community, such people can be hired, drafted, or they may volunteer, into what may be called immune teams. Immune teams will serve as support staff in nursing homes and in hospitals, possibly after undergoing rapid emergency training which will allow them to also serve as nursing aides. Then, such immune teams could replace support staff and possibly, partially, some tasks of the medical staff who have been infected or are in isolation, thus helping prevent the collapse of these systems. As their numbers grow, they will also replace susceptible personnel who have not yet contracted the disease, to further protect the population at risk from being infected. As members of the staff recover, they will return to their positions, gradually replacing the less-trained immune teams. When herd immunity is achieved, including in the support and medical staff, normal functioning can resume.

If the vulnerable population could meet only members of the immune teams and no one else, the vulnerable population would be absolutely protected by the IBS. This would lift the main burden from the health system, and will thus allow most of the world to return to normal activity. However, this is unrealistic. To assess the possible benefits of a realistic IBS, by which some of the caretakers of the vulnerable population are irreplaceable, we constructed a mathematical model which examines this strategy and performed simulations under a range of assumptions (see [8] for a review of various mathematical models of COVID-19). The mathematical model we propose is a variant of SEIR type models with several compartments and infection stages as detailed below (in particular it includes symptomatic and asymptomatic infections and allows to also examine the influence of re-infections). Importantly, despite the usual notorious sensitivity of our model to parameters such as social distancing [9], the model shows that the immune buffer strategy is robust.

We first describe the model construction (with additional details in the Appendix), then its results, and then its implications.

## 2 The Immune Buffer Strategy model

The mathematical model we derive is a variant of the SEIR models [10], see Figure 1. A traditional SEIR model describes the dynamics of a single population across 4 stages (Susceptible, Exposed, Infected and Recovered). Here, we describe the more elaborated dynamics, with both recovery or death as possible end stages, and with dividing the infected stage into three stages: those infected without symptoms, with symptoms and those in isolation. When tracking the dynamics of different sub-populations, we add compartment for each sub population, and model the interaction between those sub-populations by cross compartments interaction terms.

**Figure 1:**
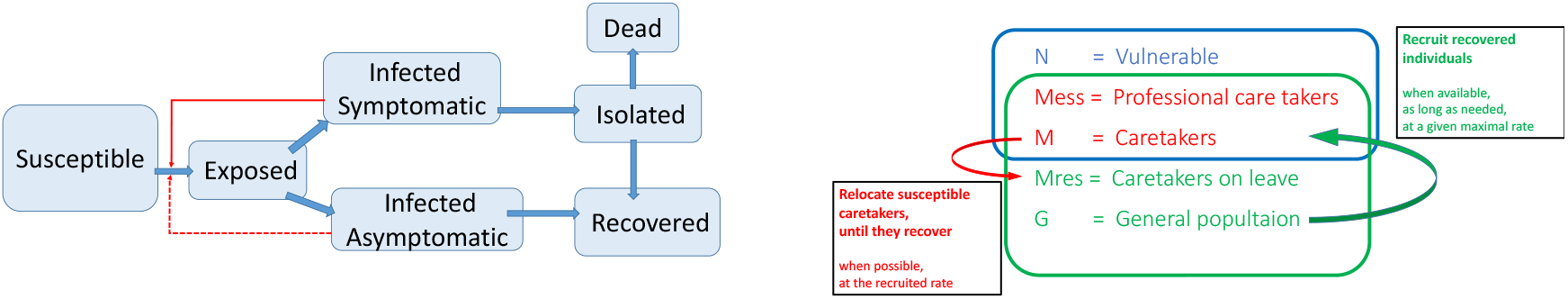
Block diagram of the mathematical model and the immune-buffer intervention. There are 5 compartments, divided to two rings of interactions. Each compartment has 7 stages, with two of them causing infections to members of the same ring. The vulnerable population interactions with each compartment is additive whereas the general population interactions with the staff is mixed.

Specifically, we divide the population into five compartments: the vulnerable individuals (denoted by *N*), the essential caretakers (*Mess*, who, due to their specialized professional skills, cannot be replaced), the non-essential caretakers (*M*, who can be replaced by immune teams), the caretakers who are on leave (denoted by *Mres*), and, finally, the general population (*G*). Each of the 5 sub-populations is further divided to 7 stages: 1-Susceptible, 2-Exposed, 3-Infected and Asymptomatic, 4-Infected and symptomatic, 5-Isolated, 6-Recovered, 7-Dead (the division to these stages is similar to the single population model of [11]). Susceptible individuals become Exposed (the latent stage of the disease) due to contacts with infected symptomatic or asymptomatic individuals. Exposed individuals who become symptomatic move to the Isolated state from which they either recover or die. Exposed individuals who become asymptomatic infect the susceptible population at a weaker rate, until they recover and join the Recovered state.

The cross-compartment interaction terms ensue from the following assumptions: the on-duty caretakers come into contact with both the general population and the vulnerable population, but, individuals from the general and from the vulnerable populations do not have direct contacts. Thus, the caretakers are the buffer between the general and the vulnerable populations. In the absence of intervention, this buffer is leaky: some of the caretakers become infected and infect vulnerable individuals who in turn infect additional vulnerable individuals and caretakers. The intervention is aiming to reduce this leakage. Currently, these chains of infections can cause devastating outbreaks leading to deaths in vulnerable communities (see e.g. [12]). The governments and the public monitor such outbreaks and employ social distancing strategies that lower/raise *R*_0_ as well as the extra precautions taken when dealing with the vulnerable population. The dashed line in Figure 2 shows such outbreaks in our model with no intervention (*R*_0_ and the vulnerable-caretakers protection factor are changed at policy changing dates, see below).

**Figure 2:**
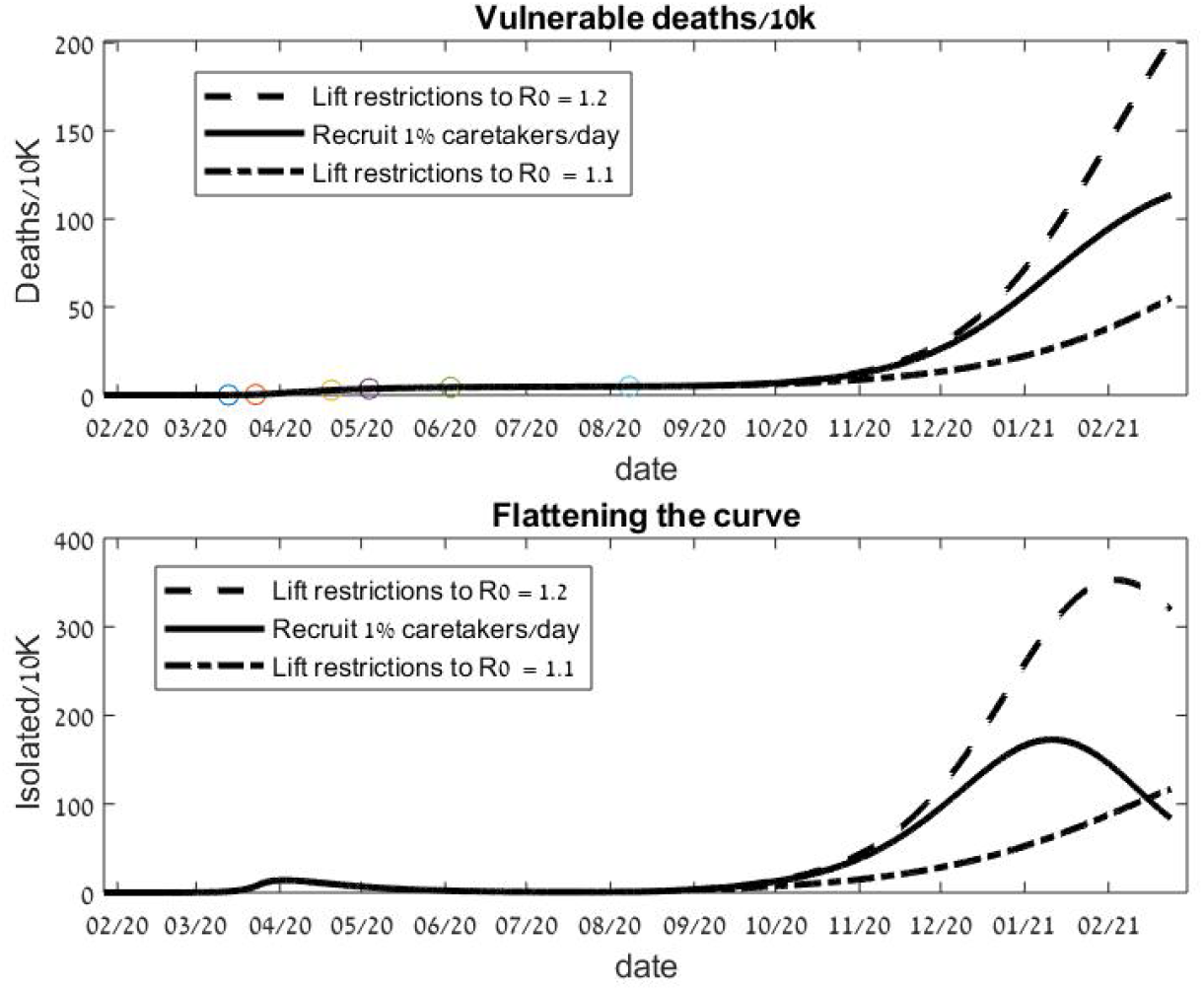
Death toll reduction and flattening of the curve by the immune-buffer strategy. In a hypothetical strategy for Germany, using parameters that fit the Germans’ historical data till August 8, 3 exit strategies are compared: 1) *R*_0_ kept at 1.2 and restrictions on interactions of the care takers and the vulnerable population are lifted to *µ_M_* = 0.5 (dashed line) 2) Under the same conditions employ the IBS with recruitment rate of 1%/day of the caretakers from the recovered population (solid line). 3) Restrict the general population so that *R*_0_ is reduced to 1.1 with *µ_M_* = 0.5 (dotted line).

Employing the IBS intervention, the fraction of recovered individuals in the caretaker compartment is increased through recruitment of recovered individuals from the general population (immune teams), at a maximal set rate, pending on availability. In Figs 2,3, this maximal rate is set to be near the optimal rate of 1%/day of the initial caretakers, *M*(0). Simultaneously, the number of potentially infectious caretakers is decreased by the same rate, when possible, by moving susceptible non-essential caretakers to the susceptible on-leave caretakers compartment, *Mres*. In addition, when susceptible caretakers (either those still working or those on leave) become infected and recover, they return to work, replacing the immune teams which had been recruited from the general population.

**Figure 3:**
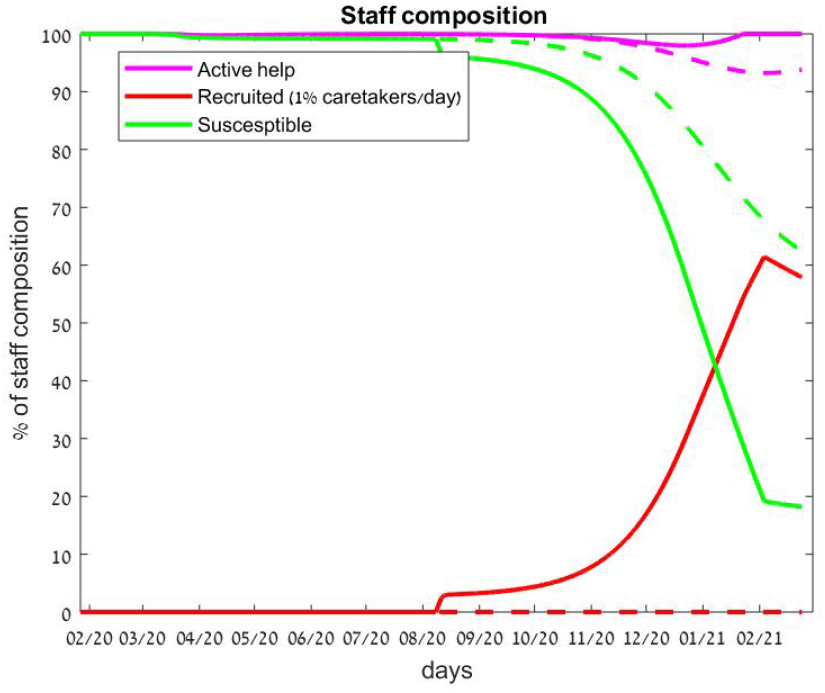
The immune buffer strategy caretakers composition. A recruitment (solid red) of 1% per day of the caretakers and a release of susceptible caretakers to other tasks lowers the fraction of the susceptible staff (solid green) while maintaining the needed staff (solid magenta). The corresponding fractions of the the needed staff and the susceptible staff (of both the essential and non-essential caretakers) without the intervention are shown by the corresponding dashed lines.

The parameters (rates of transfer among the different stages and the parameters for a-symptomatic infections) are taken, as far as possible, to be as in the Imperial-UK modelling study [12] (see Table 1). The additional parameters in our model (e.g. the duration of the isolation stage and the lower infection rates of the vulnerable compartment within itself and with the caretakers as well as the death rates) are set to reasonable values which keep the results without intervention consistent with the available data and allow fitting. Briefly (see details in the Appendix), calibration was done first to the Israel data set of active cases and deaths [13] (active cases assumed to reflect symptomatic cases due to the test strategy of Israel). Seven fitting parameters were optimized by a least square fit to the data: *R*_0_ at each date of the 6 policy changes in Israel till August 2, and the non-essential help reduction factor *µ_M_* that was changed at the lock-down date from its initial value to the fitted value (without this change the number of deaths did not match the data). To examine the IBS effectiveness, this parameter, *µ_M_*, is changed to *µ_M_* = 0.5 when the IBS is employed; its initial value is *µ_M_* = 0.6 and after lock down it is reduced to *µ_M_* = 0.2, reflecting the strong regulations in elder’s homes. For Germany, the help reduction factor after lock-down was taken to be the one fitted for Israel, and the 6 *R*_0_ values at the policy changing dates were fitted only to the number of deaths from the data set, as it is suspected that the number of symptomatic patients in Germany was larger than reported due to the selection protocol of individuals to be tested.

**Table 1:** Parameter values.

| Parameter name | Symbol | Value |
| --- | --- | --- |
| IBS Parameters |  |  |
| IBS $R_0$ ** | $R_0$ | 1.2 (Germany), 1.2 (IL) |
| IBS Help Red.** | $\mu_M$ | 0.5 |
| IBS date** | $T_{reg}$ | 7/8 |
| Daily recruit rate % | $Perrec$ | 1% |
| Epidemiological parameters |  |  |
| 1/Latency period* | $\gamma_{\alpha}^{EI}$ | 1/3[days] [12] |
| 1/Infectious period* | $\gamma_{\alpha}^{IIs}$ | 1/1.5[days] [12] |
| 1/A-sympt. infectious period* | $\gamma_{\alpha}^{AsR}$ | 1/1.9[days] [12] |
| 1/Recovery period | $\gamma_{\alpha}^{IsR}, \alpha \neq N$ | 1/30[days] |
| 1/Vuln. Recovery period | $\gamma_N^{IsR}$ | 1/15 |
| Vul. Death rate | $\gamma_N^{IsD}$ | 0.02/[days] |
| Death rate (non-$N$) | $\gamma_{\alpha}^{IsD}, \alpha \neq N$ | $2.66 \cdot 10^{-05}$/[days] |
| Symptomatic fraction | $\sigma$ | 0.66 [12] |
| Asymptomatic Reduction factor | $R_{As}$ | 0.66 [12] |
| Vulnerable reduction factor | $\mu_N$ | 0.05 |
| Essential Help Reduction factor | $\mu_{Mess}$ | 0.05 |
| Help Reduction factor* | $\mu_M$ | 0.6, 0.2 |
| Re-infection Reduction factor | $\rho$ | 0 |
| Policy change dates, Germany | $T_{reg}$ | 27/1 13/3, 23/3, 20/4, 4/5, 3/6 |
| Fitted $R_0$ at changed dates*, Germany | $R_0$ | 2.13, 2.16, 0.61, 0.46, 0.77, 1.2 |
| Policy change dates, Israel | $T_{reg}$ | 21/2 18/3, 1/4, 1/5, 20/5, 7/7 |
| Fitted $R_0$ at changed dates*, Israel | $R_0$ | 2.37, 1.97, 0.38, 1.13, 1.50, 0.92 |

\*The second value of $\mu_M$ is set from the lock-down date (here the third policy change date) till the IBS date.

Tables 1 and 2 provide the parameters and initialization of the model, Table 3 provides the sensitivity analysis to each of the parameters with and without the IBS, and Figures 9 and 10 show the resulting fitting to the historical data sets.

**Table 2:**
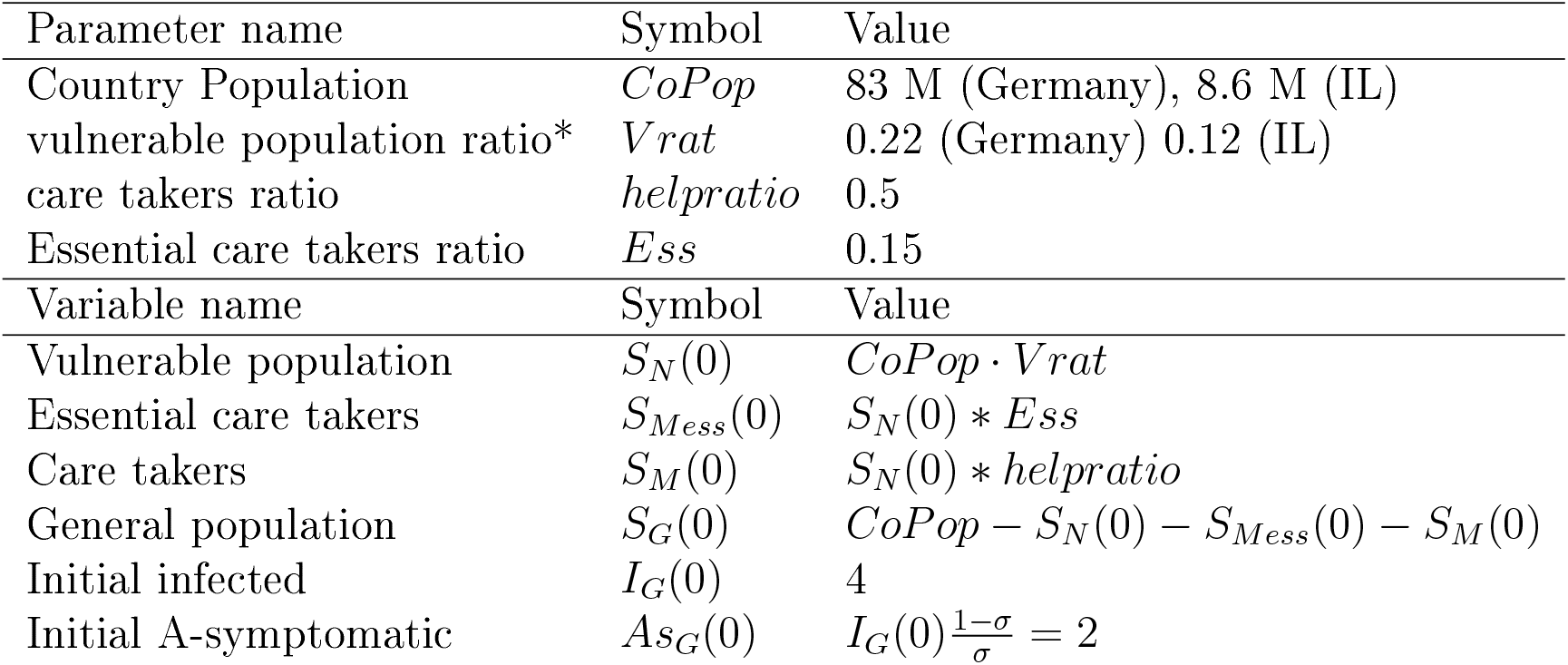
Initial setup

**Table 3:** Sensitivity to Parameters: deaths and maximal loads of isolated symptomatic vulnerable individuals (per 10K vulnerable) 6 months after employing IBS with and without employing IBS.

| Parameter name | Range | Deaths<br>Control | Deaths<br>IBS | Max Load<br>Control | Max Load<br>IBS |
| --- | --- | --- | --- | --- | --- |
| Base values | Table 1 | 201 | 113 | 352 | 169 |
| IBS parameters |  |  |  |  |  |
| IBS $R_0^{**}$ | [1, 1.4] | [13, 462] | [12, 246] | [14, 808] | [14, 454] |
| IBS Help red.** | [0.24, 0.74] | [64, 454] | [47, 228] | [131, 727] | [72, 377] |
| IBS date<sup>a</sup> | [7/8, 7/11] | [53, 53] | [41, 41] | [108, 108] | [62, 62] |
| IBS date<sup>b</sup> | [7/8, 7/11] | [249, 249] | [126, 171] | [351, 351] | [172, 246] |
| IBS date<sup>c</sup> | [7/8, 7/11] | [201, 145] | [113, 90] | [352, 347] | [172, 162] |
| Daily recruit rate % | [0.5, 1.5] | [201, 201] | [111, 127] | [352, 352] | [172, 192] |
| Sensitive parameters |  |  |  |  |  |
| Latency period* | [4.3, 5.1] | [226, 147] | [117, 98] | [338, 33]9 | [168, 170] |
| Infectious period* | [1.3, 2] | [218, 144] | [116, 96] | [352, 345] | [168, 174] |
| A-sympt infec. period* | [1.2, 2.8] | [224, 148] | [116, 98] | [351, 345] | [166, 174] |
| Vul. pop. rat.* | [0.16, 0.27] | [154, 233] | [81, 139] | [327, 371] | [124, 208] |
| Regular parameters |  |  |  |  |  |
| Recovery period | [14, 44] | [119, 247] | [59, 148] | [198, 447] | [90, 227] |
| Vul. Recovery period | [7.5, 22] | [339, 143] | [189, 81] | [313, 369] | [153, 180] |
| Vul. Death rate | [0.05, 0.16] | [98, 276] | [56, 155] | [382, 331] | [186, 162] |
| Sympt. fraction | [0.31, 0.97] | [99, 295] | [48, 185] | [157, 530] | [65, 295] |
| A-sympt red. fac. | [0.31, 0.97] | [208, 196] | [115, 112] | [353, 352] | [171, 173] |
| Vulnerable reduction factor | [0.025, 0.075] | [192, 210] | [109, 118] | [341, 364] | [166, 178] |
| Ess. Help reduction | [0.025, 0.075] | [185, 217] | [104, 123] | [371, 371] | [160, 185] |
| Help reduction<sup>d</sup> | [0.1, 0.3] | [192, 213] | [110, 117] | [353, 352] | [172, 172] |
| Help ratio | [0.24, 0.74] | [135, 247] | [64, 154] | [311, 383] | [95, 230] |
| Ess. ratio | [0.075, 0.225] | [199, 203] | [111, 115] | [351, 354] | [169, 175] |
| Init. sympt. infections | [2, 6] | [166, 215] | [106, 113] | [362, 343] | [184, 161] |

\* Sensitive parameters for fitting. \*\* Sensitive parameters for policy <sup>a</sup> Fixed Help reduction factor at $\mu_M = 0.2$ , <sup>b</sup> Fixed Help reduction factor at $\mu_M = 0.5$ . <sup>c</sup> Help reduction factor increased at IBS, as in Table 1, <sup>d</sup> Help reduction, $\mu_M$ , after lock down and before IBS.

## 3 Results of the immune buffer approach model

Typical results of the fitted model are shown in Figure 2. We see that the immune buffer approach allows to relax the social distancing restrictions as it contributes to a significant reduction in the death toll and towards flattening of the curve; hereafter, a significant reduction of the peak of symptomatic sick people from the vulnerable population. At the beginning of August 2020, the value of *R*_0_ = 1.2 gave the best fit for the death data set, and lead to the beginning of a second infection wave. In Fig 2 we examine the possibility of not imposing new restrictions and even relaxing the restrictions of the care-takers-vulnerable interactions (so *R*_0_ = 1.2 and *µ_M_* = 0.5) while employing the IBS. The number of deaths with the IBS is reduced by 45% when compared to the number of deaths occurring with lifting the regulations without any additional measures. The reduction is similar to the reduction achieved by imposing further restrictions on the general population so that *R*_0_ is decreased to *R*_0_ = 1.1 (with *µ_M_* = 0.5). In fact, a longer integration of additional 100 days shows that the number of deaths for this latter case of *R*_0_ = 1.1 is larger than the one achieved by the IBS with *R*_0_ = 1.2 (see Figure 11 in the Appendix).

The novel feature of the model is demonstrated in Figure 3. By actively increasing the number of recovered individuals in the caretaker staff through recruitment of recovered individuals from the general population while, simultaneously, actively lowering the number of potentially infectious individuals (by moving, when possible, susceptible caretakers to alternative temporary employment), the caretaker staff buffer becomes gradually more immune and the idealized immune buffer limit is approached **long before the general population gains herd immunity**. Indeed, notice that the replacement of susceptible caretakers (large drop in the solid green line) is completed about a month before the large drop in the control population of the susceptible caretakers (dashed green line). This latter drop corresponds to the dangerous stage by which many caretakers get infected as the infection becomes widely spread in the general population. The daily recruitment rate of 1% of the replaceable caretakers corresponds, roughly, to the optimal recruiting rates. Smaller rates (0.5%) lead to reduced effectiveness, while higher rates hardly improve the effectiveness of the strategy (see Table 3).

The strategy is effective for other countries. Fig 4 illustrates that small countries with young population, such as Israel, can gain a reduction of above 70% in deaths and of flattening the curve by employing the immune buffer strategy. Similarly, the strategy can also be applied to smaller communities of elders, where it may be easier to recruit immune staff members.

**Figure 4:**
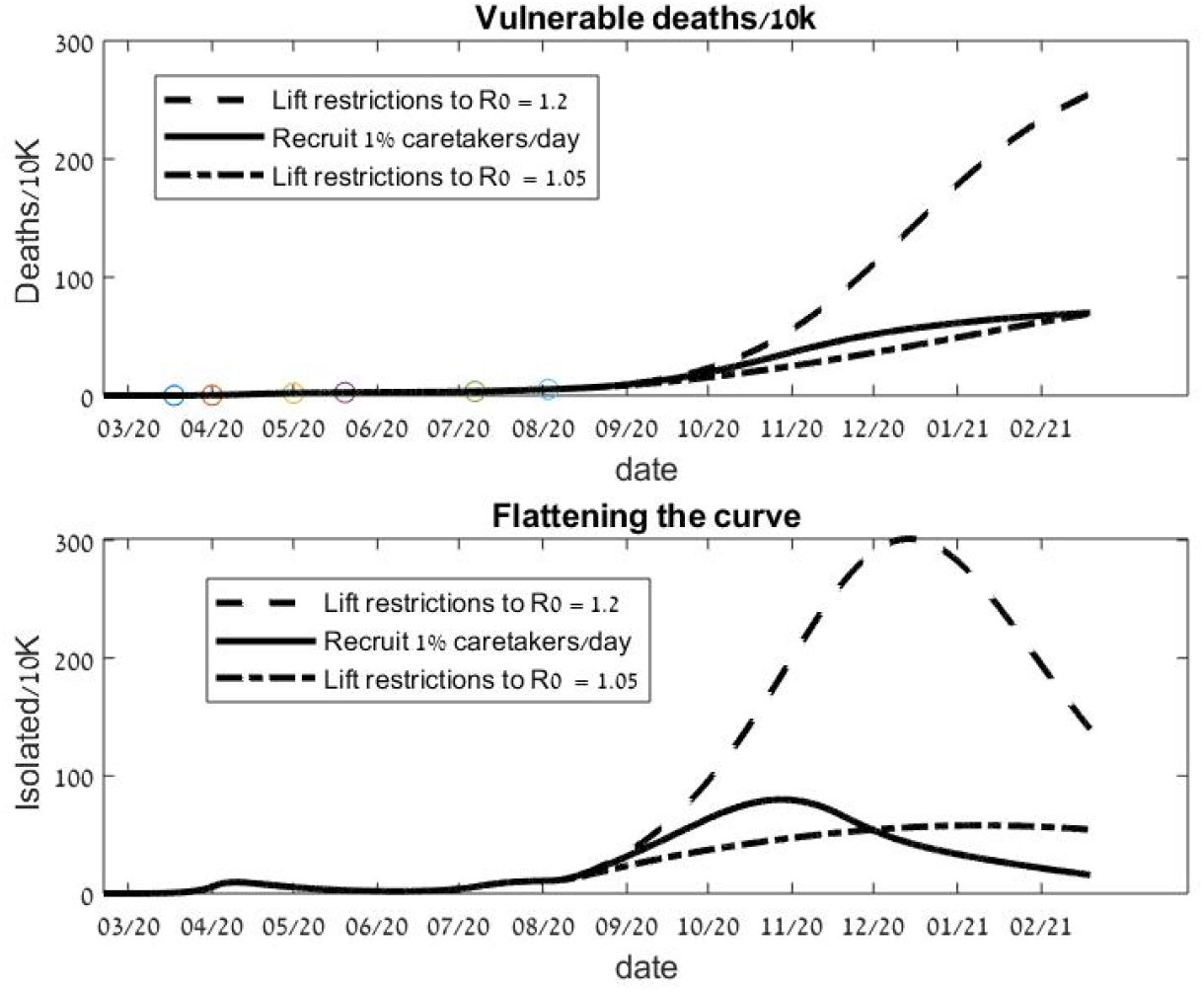
The immune-buffer exit strategy applied to a small young country. A hypothetical strategy for Israel using parameters that fit the Israeli historical data till August 2. If on August 2 the restriction are lifted to *R*_0_ = 1.2*, µ_M_* = 0.5, the immune-buffer strategy reduces deaths and flattens the curve by 70% when compared with lifting the regulation without any additional measures. For a 6 months outlook, this exit strategy is comparable to retaining restriction at the much lower *R*_0_ = 1.05.

The possibility of re-infection of recovered patients is highly debated and is raised repeatedly as a concern in general, and even more so for strategies that rely on the immunity of the recovered population. Allowing the recovered to be re-infected with a 10% rate relative to the susceptible infection rates (which is by several orders of magnitude larger than the currently known re-infection rate) shows hardly any difference in the results, see Fig 5. The same finding holds when back-infection is introduced for the simulations in Fig 4 (in [11] a different mathematical model exhibits similar insensitivity to re-infections).

**Figure 5:**
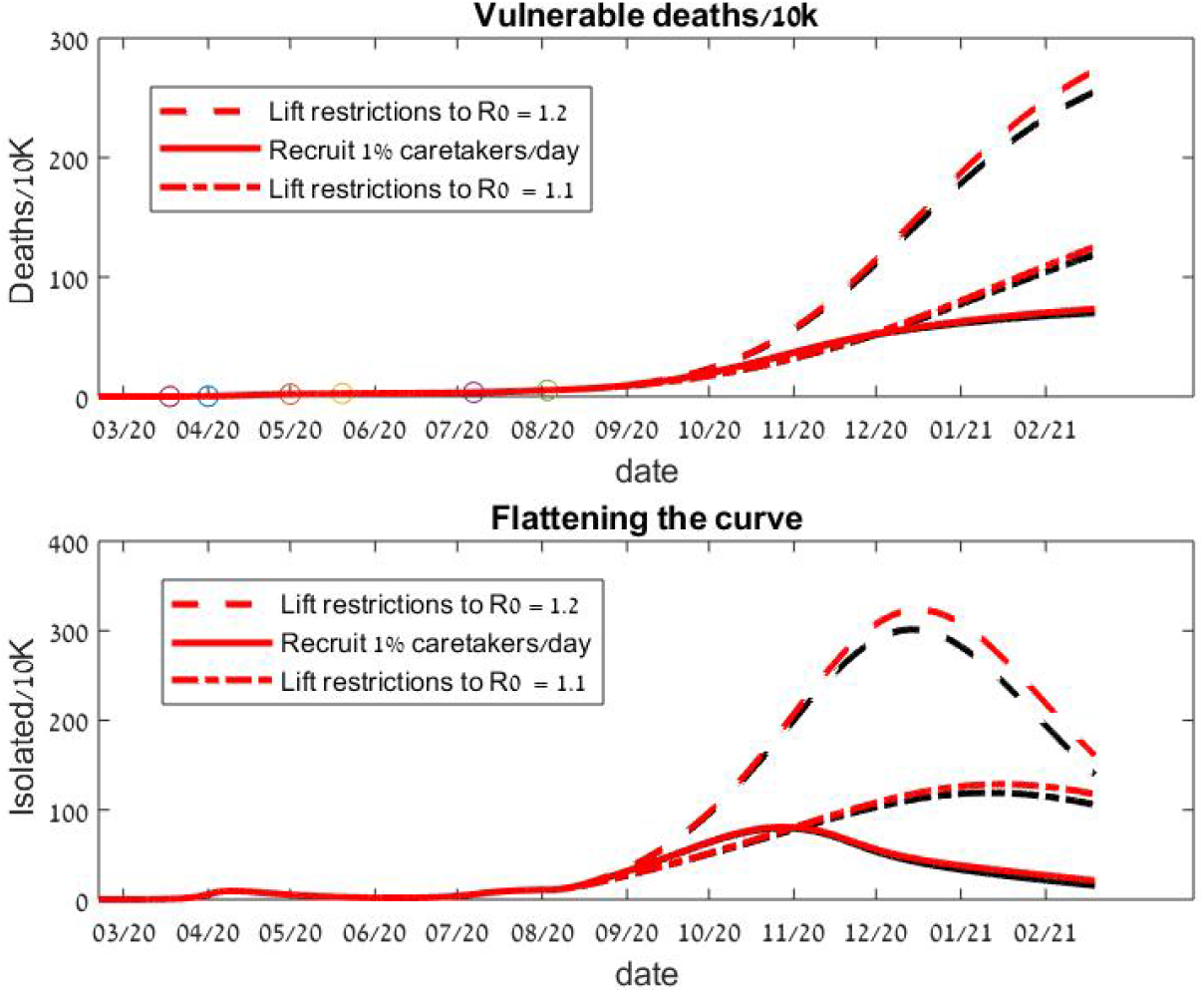
The immune-buffer strategy with hypothetical re-infections. The same conditions and strategy as in Fig 2, with possible re-infection of recovered people at 10% rate(red) and 0% (black) of the corresponding susceptible group.

More generally, the strategy is robust to changes in parameters as summarised in Table 3 and in Figures 6 and 7. In Table 3 we fix all parameters and i.c. as in Tables 1 and 2 for the Germany values (similar results are found for Israel parameter values), and change a single parameter by either ±50% or, if such a change leads to number of deaths which is either 50% below or 150% above current values, the parameter is defined as a sensitive parameter ^1^. In such a case the range is set so that the number of deaths will be in this range. The resulting variation in the control and in the Immune-Buffer Strategy deaths and maximal loads per 10K of the vulnerable population along these intervals are recorded (to gain intuition, the ordering at the intervals end points is kept).

**Figure 6:**
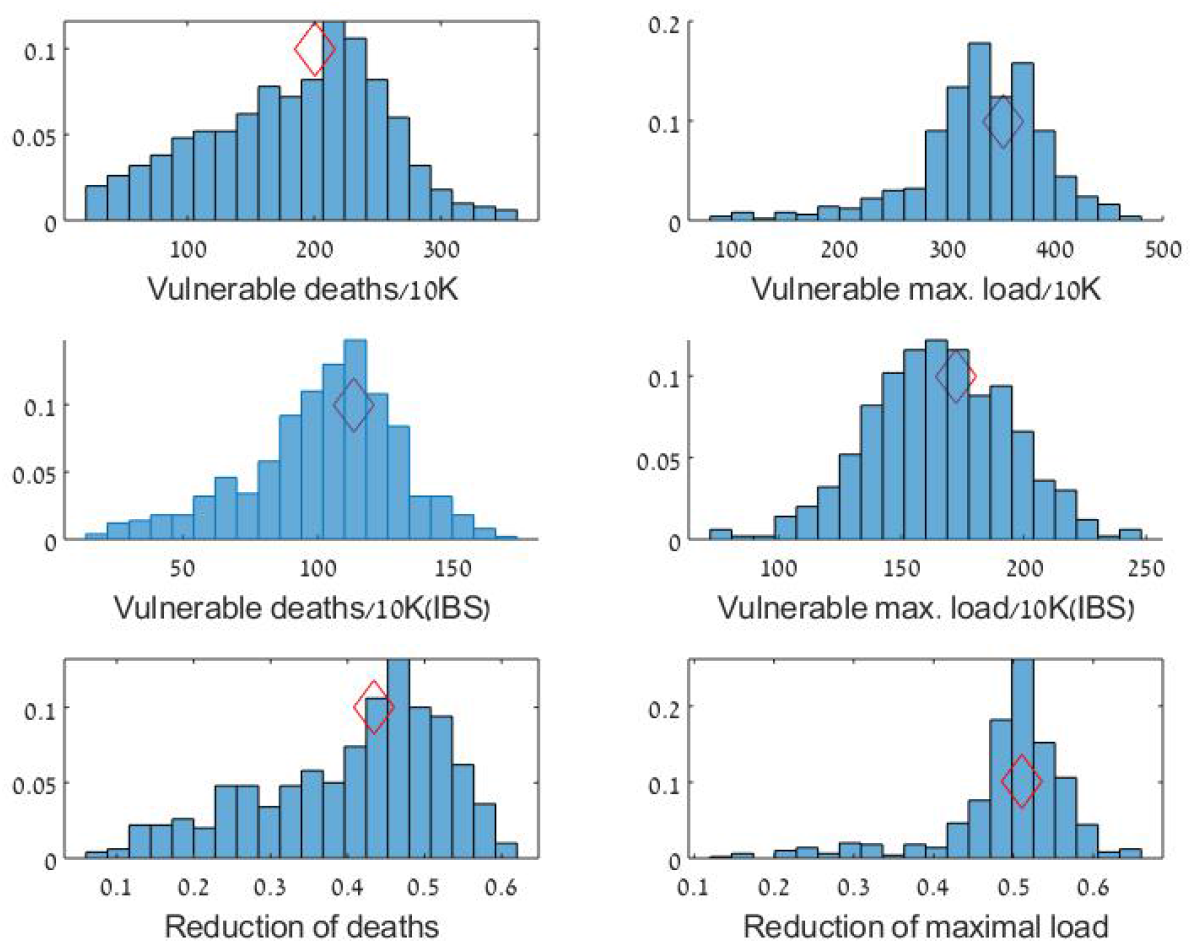
Robustness of the immune-buffer strategy (IBS), Germany. The distributions of A) Control vulnerable deaths, B) Control vulnerable maximal loads C) Vulnerable deaths with IBS strategy, D) Vulnerable maximal load with IBS E) Reduction in deaths (1-(deaths w IBS)/(control deaths)), F) Reduction in maximal loads. The distribution is achieved for 500 runs, with variations of the parameters and initial conditions as described in the text. The results are presented per 10K of vulnerable individuals. The mean,median and the coefficient of variation of the distributions are: A)(*mean, median, cv*) = (183.9, 195.2, 0.38) B) (330.3, 335.6, 0.18) C)(165.2, 165.6, 0.17) D)(101.8, 106.3, 0.0.27) E)(0.4, 0.43, 0.29),F(0.49, 0.5, 0.16))

**Figure 7:**
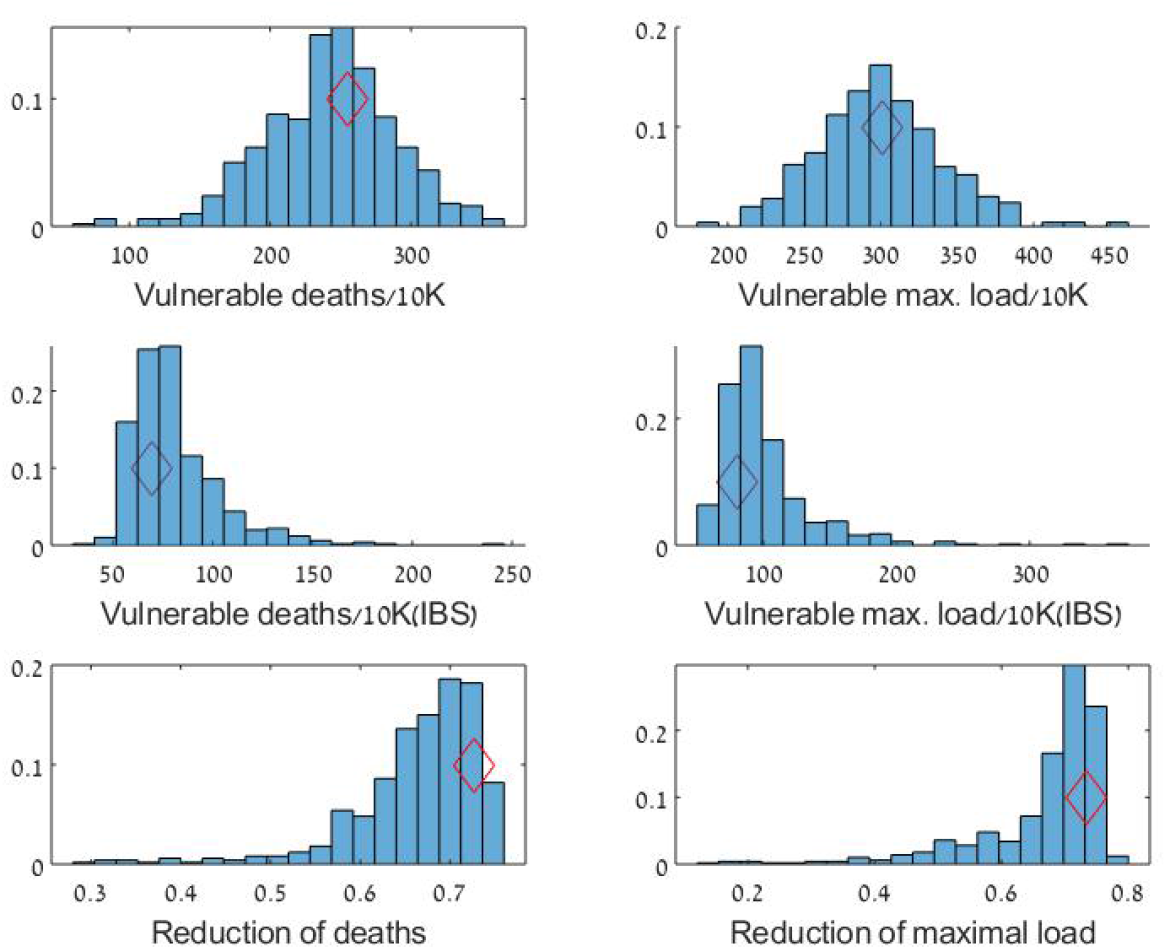
Robustness of the immune-buffer strategy (IBS), Israel. Same as Figure 6 with parameter mean values set for Israel. A)(*mean, median, cv*) = (242.5, 244.9, 0.19) B) (299.5, 296.9, 0.13) C)(99.7, 89.7, 0.35) D)(80.7, 75.5, 0.27) >E)(0.66, 0.68, 0.11),F(0.66, 0.7, 0.15))

Table 3 shows that for a fixed parameter set, with a single parameter being varied, for most parameters, a three fold change in this single parameter leaves the IBS effective in cutting by approximately half the deaths and maximal loads when these are higher than current values. Notably, for many of these (e.g. the symptomatic fraction), such a three fold change can change the number of deaths and/or the maximal loads considerably (for the symptomatic fraction – a three fold change). Yet, even though the evolution of the epidemic depends significantly on these parameters the IBS effectiveness does not: the cases where the IBS effectiveness is decreased are only those with significantly low death numbers. In particular, when the IBS help reduction factor *µM* is too low (e.g. kept at the fitted value of 0.2) the number of deaths remains low and the IBS is not needed and is ineffective.

Interestingly, the dynamics without intervention is sensitive only to the following parameters: the symptomatic and asymptomatic infection periods, latency period and the fraction of the vulnerable population (and, clearly, to *R*_0_ and *µ_M_* – the dependence on the *R*_0_ parameters is shown after the IBS policy change as, prior to the IBS employment, these parameters are used as fitting parameters for the prior data).

In Figure 6 (respectively Fig 7), histograms of 500 runs with roughly ±10% variations in all regular parameters and ±2% variations in the sensitive parameters for the Germany parameters (respectively the Israel parameters) are shown. More precisely, to retain the positive character of the parameters and initialization, we draw each parameter *p* with mean 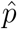 and standard deviation 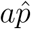 from a the Gamma distribution 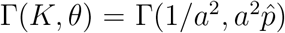 where *a* = 0.05 for regular parameters and *a* = 0.01 for the sensitive parameters (the mean 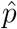 is taken from Tables 1 and 2 and the sensitivity is determined from Table 3, similar results are found for uniform distribution of the parameters on the intervals 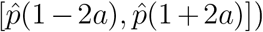. Comparing the first two rows of Figure 6 we see that the mean of the distributions for the number of deaths and for the maximal loads are reduced by more than 45% and 60% respectively for Germany (and by more than 65% and 70% for Israel, see Figure 7).

These figures (and Table 3) suggest that the strategy moves the right part of the deaths/maximal load distribution to the left, namely that it is most effective for the critical situations where the number of casualties is large. In Figure 8 we draw the correlations between the deaths /maximal loads and their reductions. For low deaths values, the IBS effectiveness is correlated with the number of deaths and the maximal load. For example, the Pearson correlation coefficients between the IBS effectiveness and the control vulnerable deaths for the 500 runs of Figures 6 and 7 are p = 0.89 for Germany (red), and p = 0.4 for Israel (Blue). Notice that there are hardly any blue data points with vulnerable accumulative deaths below 200/10K, explaining the lower *p* value. For larger values of deaths, the IBS is effective and reduces deaths by 45%-75% and maximal loads by 45%-80%, but correlation is lost and other factors (e.g. the population demographic structure) play a role.

**Figure 8:**
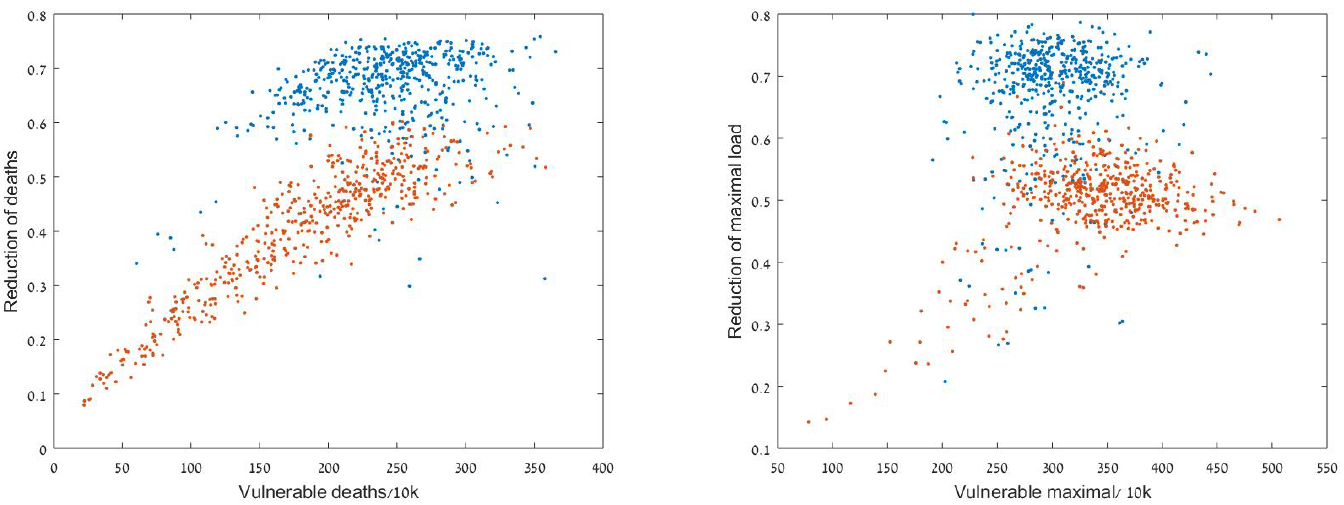
The immune-buffer strategy effectiveness increases with the level of the crisis. Cross correlations for Figures 6 (red) and 7 (blue).

## 4 Discussion

Our mathematical model includes modeling the balance between the restrictions imposed on the general population and those imposed on the vulnerable population which is needed for protecting the vulnerable population. If perfect sterility measures of the caretakers could be employed, the vulnerable population would be shielded from infections and the restrictions on the general population could be less tight. Since perfect sterility is impossible to achieve when the vulnerable person needs personal help, and, even when the vulnerable individuals are physically independent, isolation has negative medical, sociological and psychological effects, a balance between the global and local restrictions is needed. The mathematical model demonstrates that both these restrictions can be somewhat relaxed by employing the IBS which provides an effective middle way as demonstrated in Figures 2–7. Indeed, implementing the immune-buffer strategy is expected to result in significant reduction in the number of deaths in the vulnerable population and in ‘flattening the curve’ of incidences – the curve of infected vulnerable individuals. As shown in Figures 2 and 4, in realistic scenarios (e.g. for parameters that reasonably fit the available COVID-19 German and Israel data sets, respectively), the strategy can reduce the number of deaths and reduce peak incidence by 35%*−*70%. Namely, in the presence of this intervention, social distancing restrictions on the general population can be somewhat eased without increasing the burden of the disease in the vulnerable population. The strategy remains effective under a variety of conditions, including scaling to smaller vulnerable populations (such as nursing home networks), various hypothetical values of asymptomatic infection, and even allowing for a small rate of re-infection. On the other hand, while the IBS formally works even at larger values of *R*_0_ (e.g. setting *R*_0_ = 1.4*, µ_M_* = 0.5 for the IBS values in Table 3), the number of resulting deaths quickly becomes unreasonable (even though the IBS cuts it by approximately half). So the IBS can be utilized together with a somewhat relaxed social distancing program but cannot replace it altogether.

The immune-buffer strategy may be needed for quite a long period: even when COVID-19 vaccines will be available, vaccination of the elderly may be less effective [14] or unsafe [15]. When vaccination will be available, the IBS can be implemented together with prioritization of the susceptible caretakers to vaccination.

The degree of effectiveness of the strategy depends on many parameters listed in Tables 1 and 2, which we divide to three categories: epidemiological, global interaction and local interaction parameters:

- **Epidemiological parameters** of COVID-19 are uncontrolled parameters that are determined by the virus strain and environmental factors. These parameters govern the duration of latency, infectiousness, recovery, the basic transmission rates, the percentage of asymptomatic infections and the death rates (assuming a reasonable health system). Rough estimates for the mean rates appear in several previous works (e.g., see [12]). As data is gathered, these estimates and their distributions is improved. In our model the epidemiological parameters are taken as much as possible in accordance with available estimates on COVID-19 [12]. We show that these, together with tuning of *R*_0_, provide a reasonable fit to current data, see Tables 1, 2 and Figures 9 and 10 in the Appendix. Notably, even though the infection dynamics depends sensitively on some of these parameters (see Table 3), we showed that the IBS is significant in all cases in which significant epidemics develops.
- **Global interaction parameters** are parameters that are controlled by policy – by government regulations and the public compliance with these regulations. These include the number of daily tests performed, the selection of individuals to be tested, contact tracing and isolation policies, all of which influence the infection rates and therefore the reproductive number *R*_0_. These also include additional factors such as the number of incoming infectious individuals from other countries, the amount of testing for antibodies, and the future vaccinations strategies. The various global scale restrictions are needed to protect the vulnerable and prevent the collapse of the health system. However, these global restrictions have negative medical, economical, political, sociological and psychological effects on the society. Exit strategies are needed so that such restrictions can be minimized.
- **Local interaction parameters** are parameters controlled by institutions entrusted with the care for the vulnerable and by the vulnerable individuals. They include the social distancing and hygienic practices within the vulnerable population and with their caretakers as well as the number of care takers that interact with the vulnerable population (the parameters *µ_N,M,Mess_* and *helpratio, Ess*).

**Figure 9:**
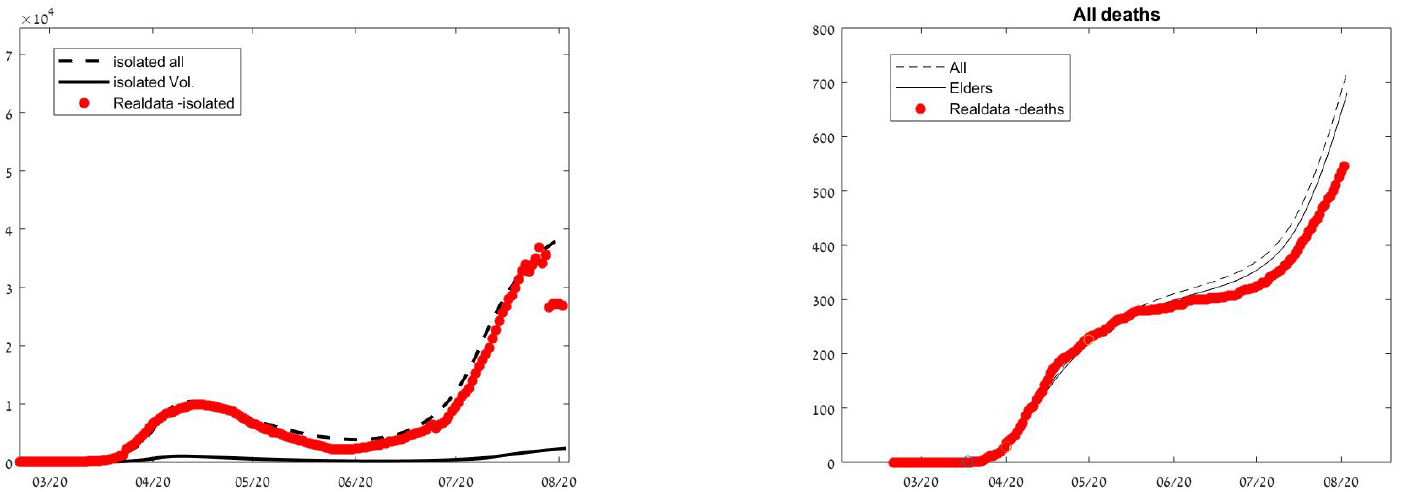
Calibration of the model for Israel: data set and a 7 parameters fitted model for both the number of deaths and the active cases (6 *R*_0_ at policy change dates and *µ_M_* after lock down).

**Figure 10:**
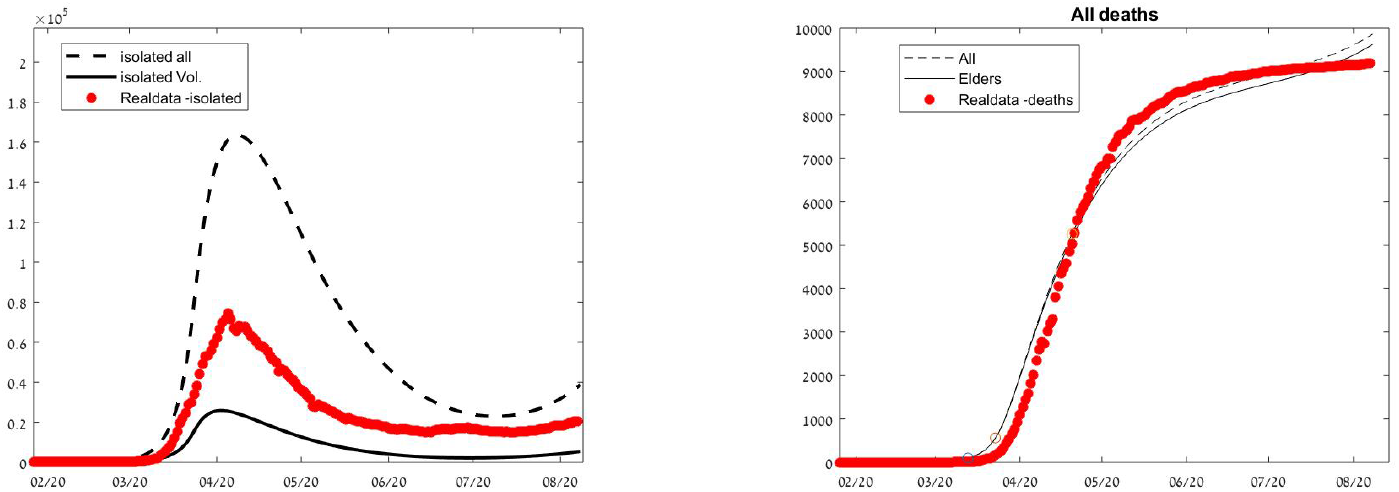
Calibration of the model for Germany: data set and a 6 parameters fitted model to the death data (6 *R*0 at policy change dates).

Clearly, both global and local interaction parameters can be altered by human actions. Yet, controlling the local interaction parameters is easier. Indeed, social distancing and enhanced sterile conditions at elders home environments are already widely employed, leading, on one hand, to less deaths than at the first outbreaks, yet, many times, to feelings of isolation and despair. The immune buffer approach allows to relax some of these measures by increasing the beneficial human contact with immunized individuals (that, for example, could use minimal protective gear). Our mathematical model (or a stochastic version of it for small vulnerable institutes) may be optimized to find what would be a reasonable protection strategy for a specific vulnerable environment at a given epidemiological and global interaction parameter values.

Implementing the life-saving immune buffer strategy can start with elders homes networks, where recruitment and hygienic regulations can be more easily implemented, it can then be extended to communities with a strong social fabric and a culture of mutual responsibility and mutual support and then to countries with effective government. Indeed, by now, all countries have rapidly accumulating cohort of individuals who are immunized to infection. This cohort includes both individuals who were diagnosed as infected and then cleared, as well as a yet-unknown portion of the population, especially among the young, who have gone through infection and recovery without being diagnosed. Means of recruiting immune teams may vary among countries and communities; 1) it may include both local volunteers and local work force teams that will be trained and receive financial compensation by government and communities (new jobs are much needed in view of the economic slowdown). 2) Countries that had an outbreak that has been contained, notably China, may send their recovered citizens as immune teams to other countries. 3) Military and national guard soldiers may be screened and deployed to assist poor vulnerable communities.

Early implementation of the immune buffer strategy makes it more effective (see e.g. the dependence on dates in Table 3). To implement it, extensive serological, PCR and other tests aimed at identifying recovered, disease free recruits from the recovered general population is needed.

Additionally, an efficient recruitment and training procedure for the recovered recruited teams and a program for alternative temporary employment for the susceptible caretakers that are relocated are needed. Such programs must also address the despotic social and ethical dangers of poor unemployed persons seeking to get infected so they can belong to the immune team.

The immune-buffer strategy aims to allow the population at risk a way to successfully cross the dire straits of the pandemic, until the safe haven of herd immunity has been reached without living in total isolation. The immune buffer strategy is not an alternative to other measures being undertaken, including social distancing and contact tracing, but it is a method which can be incorporated with other measures to better protect the population at risk, in spite of, or while tolerating, a certain level of progression of the disease among the general population.

## Data Availability

Theoretical study using public available human data.

## Acknowledgment

We thank Prof. Guy Katriel for his help in formulating the initial set up of the mathematical model and in numerous discussions along the work. VRK and OY research is supported by the ISF grant 1208/16.

## Appendix

### Mathematical model for Covid-19 with no intervention

The model equations without recruitment correspond to a seven stages model (*n*({1,…, 7}) = *{S, E, I, As, Is, R, D}*) with 5 interacting compartments *α*({1, 2…, 5}) = {*N, Mess, M, Mres, G*} (the fourth compartment, *Mres*, is empty without recruitment), where the population of each compartment (including the dead people) remains constant, see Figure 1:

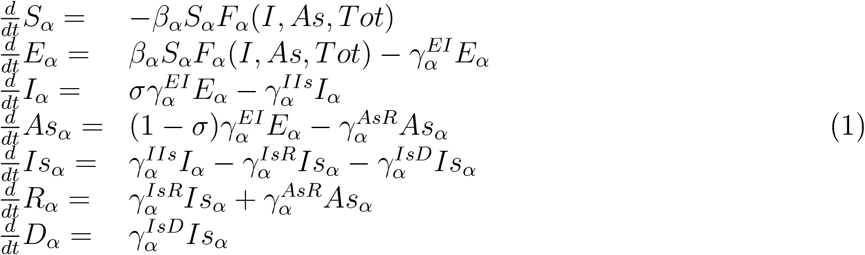

Where *T ot_α_* = *S_α_* + *E_α_* + *I_α_* + *Is_α_* + *As_α_* + *R_α_* correspond to all alive ^2^ people in compartment *α ∈ {N, M, Mess, Mres, G*} and *S;E; I;…, Tot* correspond to the vector of all compartments at the corresponding stages (here we concentrate on care-homes that transfer COVID-19 patients to a hospital. For studying hospitals' dynamics one could possibly take into account the possible infections of the staff by the isolated hospitalized patients, yet, with protection, this seems to be a minor effect).

We define

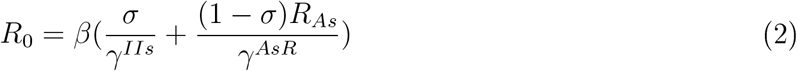

so, at each policy change we change *β* according to the new values of *R*_0_.

Denoting by 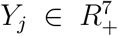 the vector of the population of all stages of compartment *α*(*j*) (so 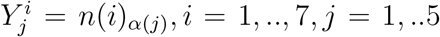, e.g., 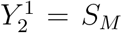 the above equations are of a block diagonal form of a linear transfer between the states and a single term *B_j_*(*y*) which corresponds to the infectious component which mixes between the different compartments. More conveniently, we re-write the equations in vector form as

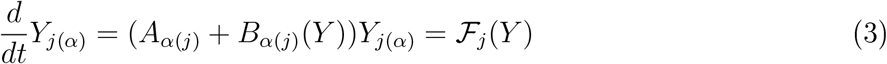

so 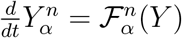, where

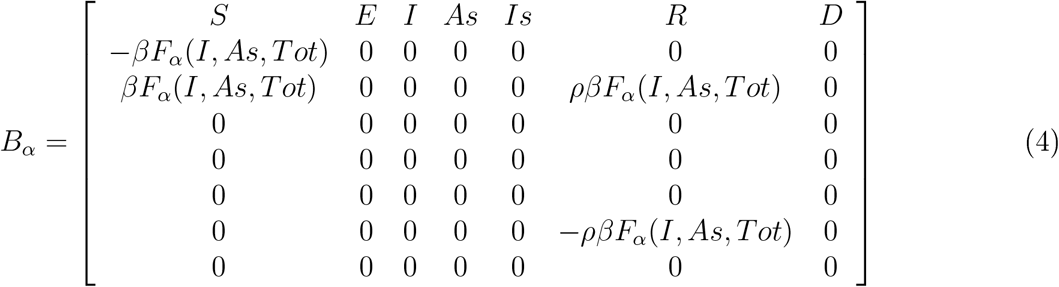

and

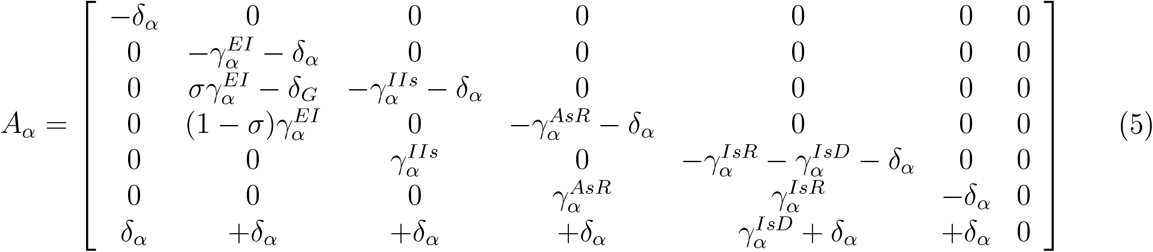

For generality sake we also include in *A_α_* the natural death term of a population at all stages, *−δ_α_* (in the simulations it is set to zero) and in *B_α_* the plausible re-infection term *ρF_α_*(*Y*) which is added to the exposed stage and subtracted from the recovered stage (*ρ* is non-zero only in Figure 5). Similarly, we can add a migration term to the general population from other communities.

The care-takers and the general population have the same internal dynamics so *A_j_* = *A*_2_*, j* = 3, 4, 5 whereas the vulnerable population has shorter recovery period and larger death rate as listed in Table 1.

With no intervention, the non-isolated persons of the first three (*N, M, Mess*) and the last four (*M, Mess, Mres, G*) compartments are in contact, where *Mres* is empty.

We assume the interactions of the vulnerable population with each compartment is separate, namely, 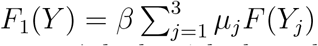 where *μ_j_* denotes the factor of the extra precautions taken when compartment *j* deals with the vulnerable population. The infection function, *F* (*Y_j_*), is the relative fraction of the two infectious stages of the *Y_j_* population (the infected and asymptomatic states), divided by the live population of the *Y_j_* population:

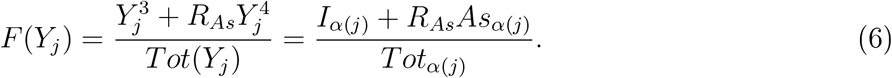

If re-infections are possible at some reduced rate, *ρ*, the recovered population can be re-infected in the same fashion as the susceptible one, as reflected by the corresponding term in the matrix *B_α_*.

We assume complete mixing between the last four compartments. Thus, the infection of the general population is 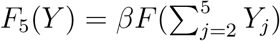 where the function *F* is defined by 6 (so *Y_j_* is replaced in the formula by the sum 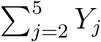). For the caretakers, the interaction occurs both with the general population and the caretakers (as for the general population) and, also, separately, with the vulnerable population, hence, *B_j_*(*Y*) = *B*_5_(*Y*) + *βµ_j_F* (*Y*_1_), where *µ_j_* corresponds to the extra protection of the *j* th caretaker compartments.

### Calibration of parameters

The model (3) depends on the following parameters; The interaction term *F_α_*(*Y*) depends on 5 parameters (*β, R_AS_, µ_N_, µ_M_, µ_Mess_*), the matrices *A_α_, j*(*α*) = 1,…, 5 depend in principle on 31 parameters 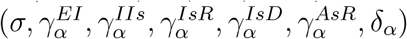, yet, with the exception of 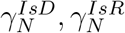, we take all the parameters to be independent of *α*. We also fix for all the simulations *δ_α_* = 0 and usually (with the exception of Fig 5) *ρ* = 0, so, finally, we have 8 free parameters for *A_α_* and all in all 13 non-trivial epidemiological parameters that are set as in Table 1. Due to policy changes by the governments, *R*_0_ and *µ_M_* change at prescribed dates at which policy changes are announced as explained next. All other parameters and initial settings are fixed as listed in Tables 1 and 2 (there, parameters were chosen as in [12] when available and by crude estimates otherwise. The sensitivity to all these parameters and initialization is listed in Table 3). In particular, all parameters that are crudely estimated are not sensitive.

### External Data

In order to validate the model, we used the daily counts of COVID-19 related deaths, and counts of positive COVID-19 tests (also referred as active cases), as reported in [13].

This data needs to be complemented by data regarding changes in government policy and in public media announcements, as these change the population behaviour – both *R*_0_ and *µ_M_*. We thus found, from Wikipedia [16], the description of the pandemic development in each country till the beginning of August. From this description we chose 6 main dates in which we believe a major change in the public behavior occurred.

### Calibration

Calibration was done first to the Israel data set of active cases and deaths till August 2nd, 2020, as shown in Figure 9 (active cases assumed to reflect symptomatic cases due to the test strategy of Israel). At the lock-down date (the third policy change date), we checked for each *µ_M_* = 0.1,…, 0.6, what is the best least square fit to the data of a vector of 6 *R*_0_ values at the 6 policy change dates. The vector of these values was chosen randomly with uniform distribution within an interval around an initial guess ([2.2 – 2.6], [1.5 – 2], [0.3 – 0.55], [0.8 – 1.2], [1.4, 1.8], [0.8 – 1.2] with best fit found for *µ_M_* = 0.2 and [2.37, 1.97, 0.38, 1.13, 1.50, 0.92]).

Figure 9 demonstrates that a good fit is achieved. Interestingly, we did not succeed to fit both the number of deaths and the number of active cases without fitting a change in *µ_M_* after lock down.

With the fitted *µ_M_* change at lock down, we fit to the German deaths data the 6 *R*_0_ values at the dates of policy changes of the German government till August 7th 2020 using the same method described above for the IL fit. The intervals for *R*_0_ were chosen to be ([1.9 – 2.3], [1.8 – 2.2], [0.3 – 0.7], [0.4 – 0.8], [0.7, 1.1], [1 – 1.4]) with best fit found for ([2.13, 2.16, 0.61, 0.46, 0.77, 1.2]). Here, the number of reported active cases is much smaller than the simulated number of cases. We chose not to fit these curves as we believe that due to the historic low-testing policy in Germany the simulations are closer to the real numbers. We see that with the increased level of testing in Germany the simulated and reported numbers become closer.

### The model with intervention

The intervention changes the model (3) as follows. We introduce in the general population compartment a recruited stage, 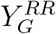, which includes, as long as needed, the recovered individuals from the general population who replace the care-takers who can be replaced. Then, the active staff becomes:

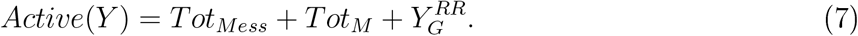

We define a flag to check whether there are enough active staff:

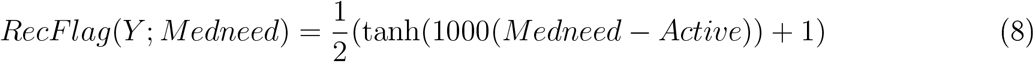

so, *RecF lag ≈* 0 when there is a surplus of staff (*Active > Medneed*) and *RecF lag ≈* 1 when recruits are needed, where *Medneed* is the total medical staff at the start *Medneed* = *S_M_*(0) + *S_Mess_*(0). We then define the following recruit and release functions:

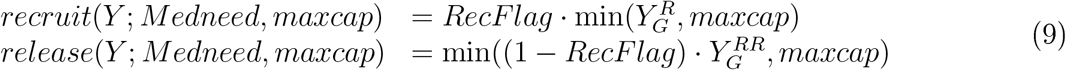

where

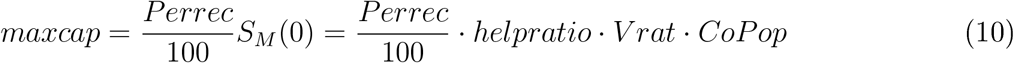

these define the recruiting and release scheme between the general population recovered and recruited stages:

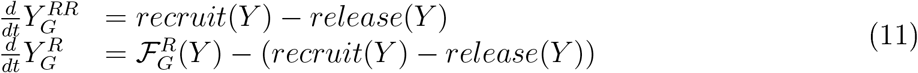

Additionally, if there are sufficient staff members in the susceptible stage (we set, for definiteness, 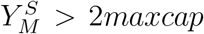) and recruitment occurs, we ask them not to attend the vulnerable population until they gain immunity, so they are moved to the *Mres* susceptible compartment. They join the active recovered staff once they recover:

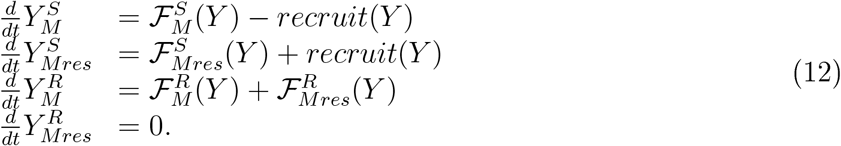

With the intervention, the infection of the vulnerable population is reduced: the term *F*_1_(*Y*) now becomes 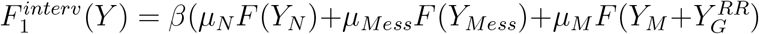 and the recovered population has no infectious states. This reduction in infections can be significant as demonstrated in Figures 2–7.

### IBS results for longer integration period

With and without the IBS, finally, the population gains herd immunity and the epidemic ends. The duration of this process and the number of casualties depend on all parameters as summarized in Table 3. In particular, these are sensitive to the changes in *R*_0_, and especially to the last IBS *R*_0_ and *µ_M_* which determine the fraction of the recovered population and the deaths. Since *R*_0_ and *µM* change with policy and public behavior, the model cannot predict such changes. Thus, in the main text we refrain from long term predictions by our model. Nonetheless, we show in Figure 11 that if one ignores such changes and keeps *R*_0_ = 1.1 for a longer period than shown in Figure 2, more casualties than in the competing IBS with *R*_0_ = 1.2 occur.

**Figure 11:**
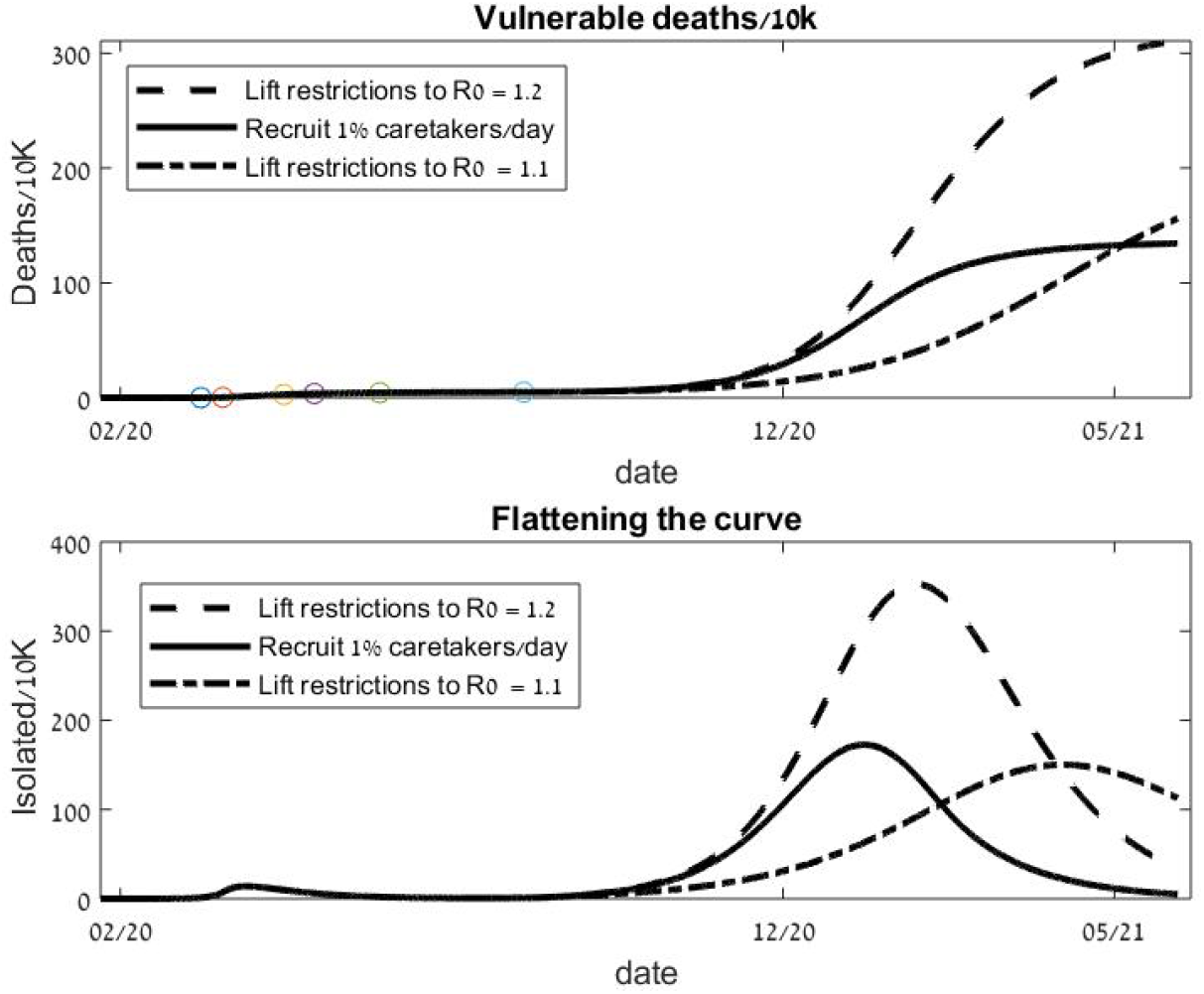
Continuation of the strategy for additional 100 days compared to 2. The IBS with *R*_0_ = 1.2 is more effective than lowering *R*_0_ to 1.1 without employing the IBS.

## Footnotes

1 We made one exception – we kept the vulnerable recovery period parameter as a regular and not as a sensitive parameter since the slightly higher death rates at the low bound (160% above the current value) reflected transient sensitivity.

2 We assume that the isolated people do not infect, yet they do reduce the number of encounters in the population.

## Notes

### Competing Interest Statement

The authors have declared no competing interest.

### Funding Statement

ISF 1208/16.

### Author Declarations

Theoretical study, no approval is needed.

## References

[1] Li, Q. et al. Early transmission dynamics in wuhan, china, of novel coronavirus infected pneumonia. New England Journal of Medicine (2020).

[2] Onder, G., Rezza, G. & Brusaferro, S. Case-fatality rate and characteristics of patients dying in relation to covid-19 in italy. Jama (2020).

[3] WHO Organization, Report of the WHO-china joint mission on coronavirus disease 2019 (covid-19) (2020).

[4] Richardson, S. et al. Presenting characteristics, comorbidities, and outcomes among 5700 patients hospitalized with covid-19 in the new york city area. JAMA (2020).

[5] Zhou, F. et al. Clinical course and risk factors for mortality of adult inpatients with covid-19 in wuhan, china: a retrospective cohort study. The Lancet (2020).

[6] Barnett, M. L. & Grabowski, D. C. Nursing homes are ground zero for covid-19 pandemic. In JAMA Health Forum, vol. 1, e200369–e200369 (American Medical Association, 2020).

[7] McMichael, T. M. et al. Epidemiology of covid-19 in a long-term care facility in king county, washington. New England Journal of Medicine (2020).

[8] Rabajante, J. F. Insights from early mathematical models of 2019-ncov acute respiratory disease (covid-19) dynamics. arXiv preprint arXiv: 2002.05296 (2020).

[9] Roda, W. C., Varughese, M. B., Han, D. & Li, M. Y. Why is it difficult to accurately predict the covid-19 epidemic? Infectious Disease Modelling (2020).

[10] Brauer, F. Mathematical epidemiology: Past, present, and future. Infectious Disease Modelling 2, 113–127 (2017).

[11] Giordano, G. et al. Modelling the covid-19 epidemic and implementation of population-wide interventions in italy. Nature Medicine 1 6 (2020).

[12] Ferguson, N. et al. Report 9: Impact of non-pharmaceutical interventions (npis) to reduce covid19 mortality and healthcare demand (2020).

[13] Johns Hopkins University. “covid-19 data repository by the center for systems science and engineering (csse) at johns hopkins university” (2020). Data retrieved from JHU CSSE COVID-19 Data on August 2 (Israel) August 7 (Germany), 2020 “https://github.com/CSSEGISandData/COVID-19/tree/master/csse_covid_19_data/csse_covid_19_time_series”.

[14] Baric, R. S. et al. Sars coronavirus vaccine development. In The Nidoviruses, 553–560 (Springer, 2006).

[15] Sharpe, H. R. et al. The early landscape of coronavirus disease 2019 vaccine development in the uk and rest of the world. Immunology 160 (2020).

[16] Wikipedia. “covid-19 pandemic in germany/israel” (2020). Accessed on September 9 2020, “https://en.wikipedia.org/wiki/COVID-19_pandemic_in_Germany”,“https://en.wikipedia.org/wiki/COVID-19_pandemic_in_Israel”.

